# Studying the Interplay Between Apolipoprotein E and Education on Cognitive Decline in Centenarians Using Bayesian Beta Regression

**DOI:** 10.1101/2020.09.25.20201962

**Authors:** Qingyan Xiang, Stacy Andersen, Thomas T Perls, Paola Sebastiani

## Abstract

Apolipoprotein E (*APOE*) is an important risk factor for cognitive decline and Alzheimer’s disease in aging individuals. Among the 3 known alleles of this gene: e2, e3, and e4, the e4 allele is associated with faster cognitive decline and increased risk for Alzheimer’s and dementia, while the e2 allele has a positive effect on longevity, and possibly on preservation of cognitive function. Education also has an important effect on cognition and longevity but the interplay between *APOE* and education is not well characterized. Previous studies of the effect of *APOE* on cognitive decline often used linear regression with the normality assumption, which may not be appropriate for analyzing bounded and skewed cognitive test scores. In this paper, we applied Bayesian beta regression to assess the association between *APOE* alleles and cognitive decline in a cohort of centenarians with longitudinal assessment of their cognitive function. The analysis confirmed the negative association between older age and cognition and the beneficial effect of education that persists even at the extreme of human lifespan in carriers of the e3 allele. In addition, the analysis showed an association between *APOE* and cognition that is modified by education. Surprisingly, an antagonistic interaction existed between higher education and *APOE* alleles, suggesting that education may reduce both positive and negative effects of this gene.

## 1 Introduction

Declines of certain cognitive abilities are common complications of aging and identifying risk factors for cognitive decline is essential to search for therapeutic interventions. Risk factors for cognitive decline include older age, lower education and genes such as apolipoprotein E (*APOE*) that plays an important role in the risk for Alzheimer’s disease (Fan et al. 2019). *APOE* is involved in the transport of cholesterol and other lipids between cellular structures (Mahley 1988). The gene has three well characterized alleles e2, e3 and e4 that are defined by the combinations of the genotypes of the single nucleotide polymorphisms rs7412 and rs429358. Studies have shown that e4-carriers have increased risk for Alzheimer’s disease and accelerated cognitive decline compared to non-carriers (Staehelin et al. 1999; Bondi et al. 2003; Wetter et al. 2005), while e2-carriers appears to have a reduced risk for age-related neurodegenerative disease (Henderson et al. 1995; Raber et al. 2004; Kim et al. 2017). The review by O’Donoghue (O’Donoghue et al. 2018) lists 40 studies of the association between *APOE* and cognition in longitudinal studies, but none of these studies examined this association among individuals at extreme ages (e.g., centenarians). Investigating the association of this gene with cognitive decline in centenarians is important for informing about the extent of the protective and deleterious effect of this gene on cognitive function at the extreme of human lifespan. In addition, examining the interaction between education and *APOE* alleles could help to better characterize the long term effect of education on cognition and the interplay between genetic and environmental risk factors. Therefore, in this work, we examined the association of the e2 and e4 alleles of *APOE* with cognitive function in centenarians enrolled in the New England Centenarian Study (NECS) (Sebastiani and Perls 2012), who are enriched for carriers of the e2 allele, have varying levels of education, and for whom we have longitudinally collected assessments of cognitive function.

Typically, studies of the association of *APOE* with cognitive decline use linear mixed models of data collected from a variety of neuropsychological tests (Blair et al. 2005; Caselli et al. 2009; Kim et al. 2017). These models assume unbounded and normally distributed errors, although the outcome of any neuropsychological test is usually defined in a limited interval between 0 and a maximum test value. In addition, the distribution of neuropsychological test scores is often skewed and asymmetric, thus introducing an additional violation of the normality assumption. Violating the normal assumption when modeling cognitive test scores could lead to a poor estimation of the genetic association with cognitive function, and predict scores that are either negative or exceed the maximum test value. To address this problem, we propose using the regression model based on the assumption that the response is beta-distributed. Beta distribution is defined in a finite interval and is flexible to accommodate distribution with different shapes. Hence this distribution is well suited to model bounded test scores and questionnaire outcomes in nature (Smithson and Verkuilen 2006).

Beta regression was proposed by Ferrari and Cribari-Neto (Ferrari and Cribari-Neto 2004) and further developed in the frequentist perspective (Smithson and Verkuilen 2006; Simas et al. 2010) and in the Bayesian perspective (Figueroa-Zúñiga et al. 2013; Cepeda-Cuervo et al. 2016). Studies in multiple fields have utilize beta regression to model different variables such as ischemic stroke lesion volume (Swearingen et al. 2011), genetic distance (Branscum et al. 2007), and understory vegetation communities (Eskelson et al. 2011). In this paper, we describe a Bayesian hierarchical beta regression model to analyze the association between *APOE* and cognitive decline among centenarians enrolled in the NECS (Sebastiani and Perls 2012).

## 2 Materials and Method

### 2.1 Participants

The NECS began in 1994 as a population-based study of centenarians living within eight towns in the Boston area and expanded enrollment to include centenarians, their siblings and offspring as well as controls to North America in 2000. Enrolled participants provide socio-demographic data, medical history, and physical function ability (Barthel Index, Mahoney and Barthel 1965; Sinoff and Ore 1997). Participants are administered the Blessed Information-Memory-Concentration (BIMC) test (Blessed et al. 1968; Kawas et al. 1995), a brief test of global cognition that can be administered over the phone to the participant or with the help of a proxy. The BIMC has a maximum total score of 37 points. Scores of 34 or greater represent no impairment, 27 to 33 indicate mild impairment, 21 to 26 signify moderate impairment, and less than 20 are associated with severe impairment (Kawas et al. 1995). NECS participants are followed annually to collect new medical events, changes in medication and physical function ability, and are administered the BIMC. The NECS study is still open for enrollment. In this analysis, we use the data collected through November 2019. Genotype data for *APOE* were inferred from the combinations of the SNPs s7412 and rs429358 as described in (Sebastiani, Gurinovich, et al. 2019).

## 2.2 Statistical analysis

### 2.2.1 Beta regression specification

A variable *y* defined in the interval (0, 1) follows the beta distribution if the density function is proportional to

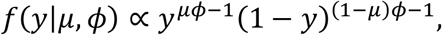

where *μ* represents the mean: 0 < *μ* < 1; the function *μ(*1 − *μ*)/(1 + *ϕ*) is the variance, and *ϕ* > 0 is the precision parameter (Ferrari and Cribari-Neto 2004). To parameterize the mean as a function of covariates, it is convenient to use the logit function

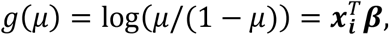

where ***β*** is a vector of coefficient and 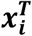 is a vector of covariates (Smithson and Verkuilen 2006; Zeileis et al. 2010). The beta distribution is defined in the open interval (0, 1). To fit this model to data defined in the range (*a, b*), we use the transformation *y*′ = (*y* − *a*)/(*b* − *a*). In our analysis, the BIMC scores range from 0 to 37 and we rescaled the data to the interval [0.01, 0.99] to avoid zeros and ones.

#### 2.2.2 Bayesian beta regression modelling

We were interested in modelling the association of the following main covariates with BIMC scores: age, sex, education and *APOE* alleles. We standardized the continuous variables *age* and *education* to generate parameters on the same scale and coded the dichotomous variable *sex* 0 for female and 1 for male. Since homozygote carriers of e2 or e4 are rare, we used the following allele groupings in the analyses:

- “e2 group” comprising carriers of the *APOE* genotypes e2e2 or e2e3;
- “e3 group” comprising carriers of the *APOE* genotype e3e3;
- “e4 group” comprising carriers of the *APOE* genotypes e3e4 or e4e4.

And we selected “e3 group” as the reference group because it is the most frequent genotype in Whites. We performed two analyses to distinguish the beneficial association of the e2 allele relative to e3 from lack of carrying the deleterious e4 allele. One analysis estimated the association between *APOE* and BIMC scores of the e2 group relative to the e3 group, including only the e2 and e3 groups. The other analysis estimated the association between *APOE* and BIMC scores of the e4 group relative to e3 group.

We used backward selection for model fitting, and started with the hierarchical regression model:

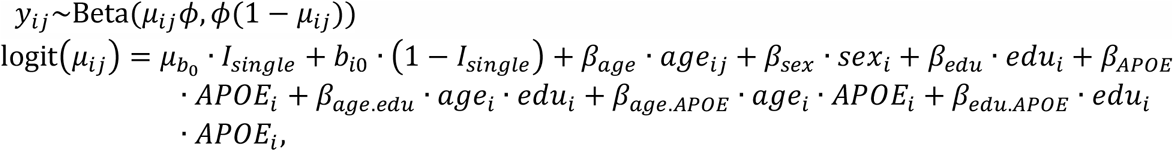

where *y*_*ij*_ denotes the *j*^*th*^ cognitive test score of the *i*^*th*^ participant, the *β* coefficients are fixed effects and *b*_*i*0_ is the random intercept that we used to account for within participant correlation of the repeated measurements. We used a piecewise random intercept *μ*_*b*0_ · *I*_*single*_ + *b*_*i*0_ · (1 − *I*_*single*_) to accommodate for participants with different number of test administration, where the indicator variable *I*_*single*_ is 0 for participants with only one cognitive test score and *I*_*single*_ = 1 otherwise. The random intercepts *b*_*i*0_ were assumed to be independent and normally distributed, i.e., 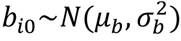. We specified a normal prior for the mean parameter *μ*_*b*_ of the random intercept that *μ*_*b*_∼*N(*0, 1000), and a gamma distribution with the shape and scale parameters both equal to 1 for the precision parameter *ϕ* and the precision of random intercept 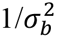. With this parameterization, the participants with only one test score would be assigned the fixed intercept *μ*_*b*_ in the regression, while participants with more than one test score have their own random intercept *b*_*i*0_. We used normal priors for all the fixed effect parameters *β* ∼*N(*0, 1000).

We implemented the backward model selection algorithm using the deviance information criterion (DIC) (Spiegelhalter et al. 2002; 2014), which is particularly useful for selection of hierarchical models. Since DIC has a tendency to overfit (Clarke and Clarke 2018), we then refined the model selected by this search by retaining only interactions and main effects with a posterior credible interval that did not include 0. Once we selected the final model, we also conducted a sensitivity analysis with respect to the prior distributions. For regression coefficients and the mean parameter of the random intercept that use the normal priors, we reduced the variance from 1000 to 10 and 100.

For precision parameter of the beta distribution and the reciprocal of the variance of the random intercept that use the gamma priors, we modified the variance of gamma priors from 1 to 100. We ran each case to assess if the parameter estimates altered after we changed the scale of the prior parameters. All analyses were conducted in R3.6 and all Bayesian models were analyzed using Markov Chain Monte Carlo (MCMC) implemented in the “rjags” package (Plummer 2016). The posterior estimates of the parameters derived from at least 8000 burn-in adaptions and 4000 iterations.

#### 2.2.3 Interpretation of the results

A limitation of beta regression is that the magnitude of the regression coefficients is not directly interpretable in terms of changes of the outcome. To better understand the association of *APOE* with BIMC scores and the interplay with education, we estimated the fitted means using inverse transformation of the logit function, and then we rescaled the fitted means to the original scale of the BIMC scores (0, 37). We calculated fitted means for the e2, e3 and e4 groups with respect to three different education levels: low education (25% quantile of study population’s education; 8 years), median education (median of study population’s education; 12 years) and high education (75% quantile of study population’s education; 15 years). For each allele in each education level, we also calculated the corresponding age of onset of moderate cognitive impairment (BIMC score = 26) using the predicted trajectories of the cognitive test scores. These ages provide a quantitatively more interpretable metric of the genetic effects.

## 3 Results

Out of 768 total participants in the NECS dataset with *APOE* genotypes, we excluded 167 participants with missing test scores, 111 participants with missing education information, and 4 participants with *APOE* genotype e2e4. Table 1 summarizes the demographic characteristics and test scores at baseline of the remaining 486 participants used in our analysis. The *APOE* e3 group was the most prevalent and used as the reference group in all subsequent analyses. Approximately 24% of centenarians in the study carried at least one e2 allele, while only 8% were carriers of an e4 allele. We did not find any e4 homozygous centenarians.

**Table 1.** Demographic characteristics and the BIMC scores at baseline of the participants in the New England Centenarian Study

|  | *APOE* e2<br>(e2e2,e2e3) | *APOE* e3<br>(e3e3) | *APOE* e4<br>(e3e4) |
| --- | --- | --- | --- |
| N | 117 | 331 | 38 |
| Age at Enrollment, mean (SD) | 103.54 ± 4.59 | 103.32 ± 4.43 | 102.42 ± 4.97 |
| Gender, male (%) | 22 (18.8%) | 85 (25.68%) | 9 (23.68%) |
| Years of Education, mean (SD) | 11.63 ± 3.51 | 11.82 ± 3.85 | 12.29 ± 4.05 |
| BIMC scores at baseline, mean (SD) | 25.11 ± 8.23 | 25.00 ± 8.63 | 24.38 ± 10.20 |

The histogram in Figure 1 shows that the distribution of the baseline BIMC scores is bounded and highly skewed. Hence we analyzed the rescaled test scores using beta regression. The combination of backward selection using the DIC criterion and refinement of the model using the posterior credible intervals produced the models summarized in Tables 2 and 3. The diagnostic plots (trace plots, autocorrelation plots and the Gelman plots of all MCMC chains) showed no indications of lack of convergence. The sensitivity analyses also showed that changing the prior distributions did not appreciably alter the parameter estimates. The diagnostic plots and sensitivity analyses are all included in the supplementary files.

**Table 2.** Parameter estimates and 95% credible intervals from the analysis of the BIMC scores in carriers of *APOE* e2 and e3 Alleles

|  | Estimates | 2.5%CI | 97.5%CI | SD |
| --- | --- | --- | --- | --- |
| Intercept ($\mu_b$) | **0.592** | **0.489** | **0.695** | **0.053** |
| Age | **-0.110** | **-0.138** | **-0.099** | **0.009** |
| Sex, male | **0.262** | **0.076** | **0.447** | **0.096** |
| Education | **0.062** | **0.039** | **0.085** | **0.012** |
| *APOE2* | 0.037 | -0.162 | 0.186 | 0.088 |
| *APOE2* *edu | **-0.063** | **-0.109** | **-0.019** | **0.023** |

All covariates were standardized and the score was rescaled to the interval (0, 1) to fit the beta regression, so all parameters should be interpreted on the logit scale and the effects are relative to the rescaled score.

**Table 3.**
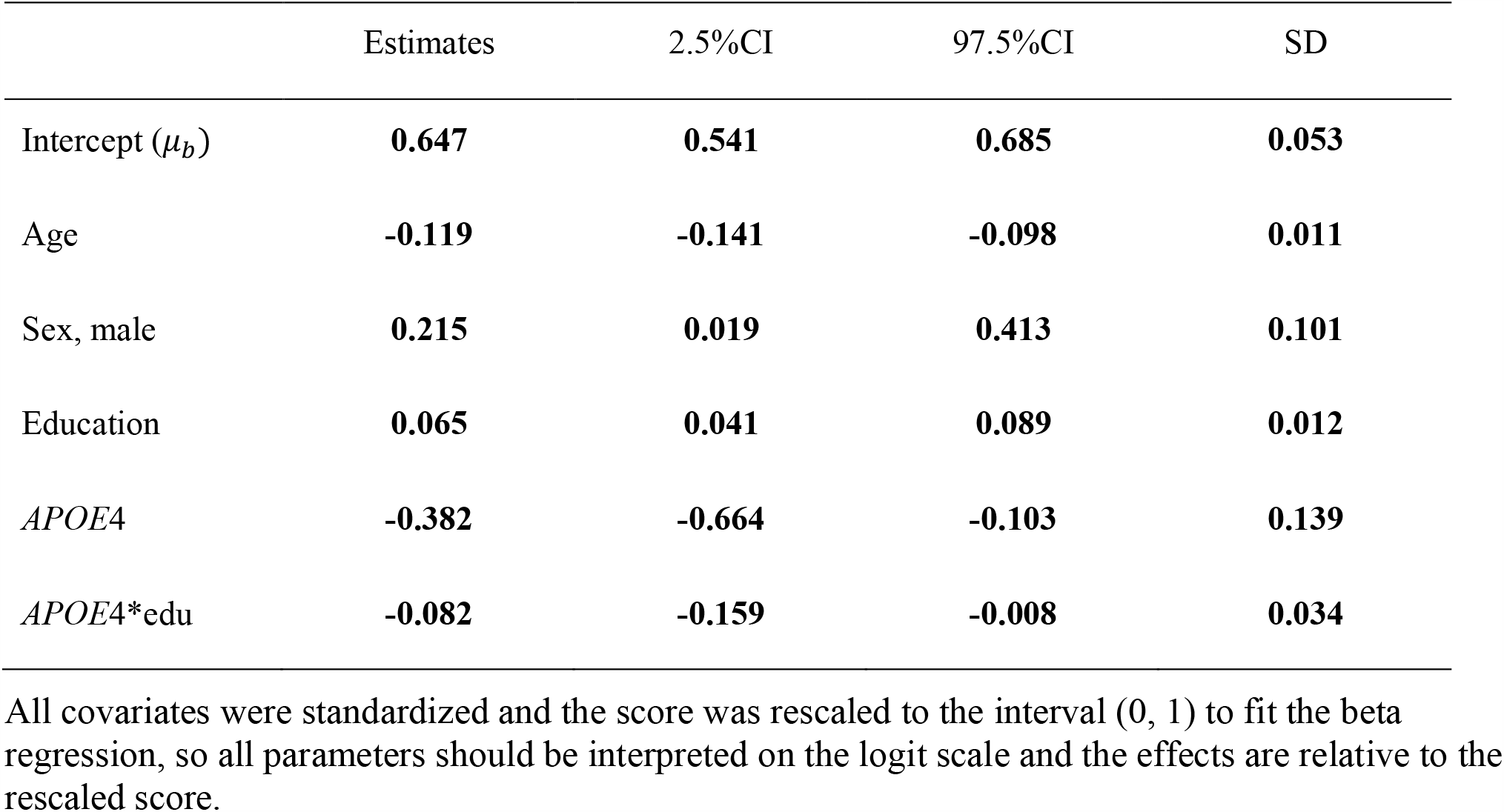
Parameter estimates and 95% credible intervals from the analysis of the BIMC scores in carriers of *APOE* e4 and e3 Alleles

**Figure 1.**
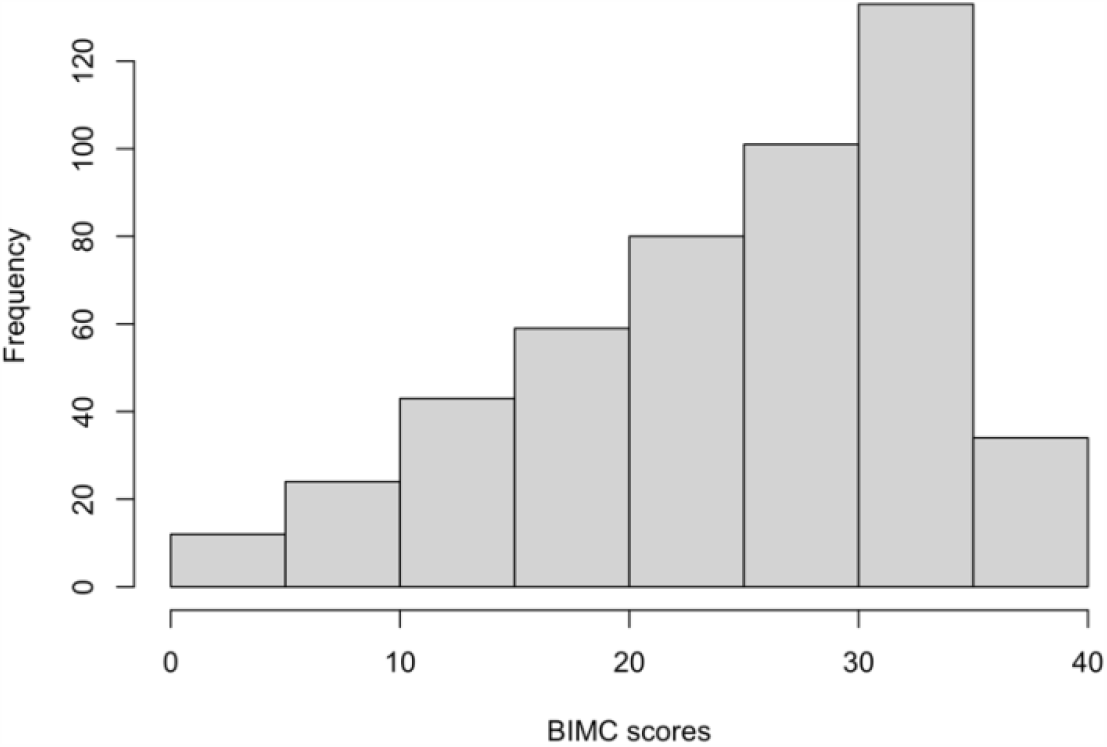
Histogram of baseline Blessed Information-Memory-Concentration scores from the New England Centenarian Study.

Table 2 summarizes the results of the analysis comparing *APOE* e2 with *APOE* e3, while Table 3 summarizes the results of the analysis comparing *APOE* e4 with *APOE* e3. The significant coefficients are marked with bold fonts. In both analyses, age, sex and education were all significantly associated with BIMC scores. Older age was associated with significantly worse performance on the test. More years of education and male sex were associated with significantly higher scores.

In the analysis comparing e2 with e3, the interaction between education and e2 was significant, thus suggesting an effect modification of education on the association of e2 with BIMC scores. It is noteworthy that the estimates of the main effect of education (0.063) and the interaction term (−0.062) were opposite, so that the negative interaction between e2 and education numerically canceled out the positive association of education with BIMC scores in carriers of the e2 allele. This is shown in Figure 2, in which the fitted BIMC scores in e2 carriers remain nearly the same in different education levels. Figure 2 also shows the positive association of education with BIMC score in the e3 group.

**Figure 2.**
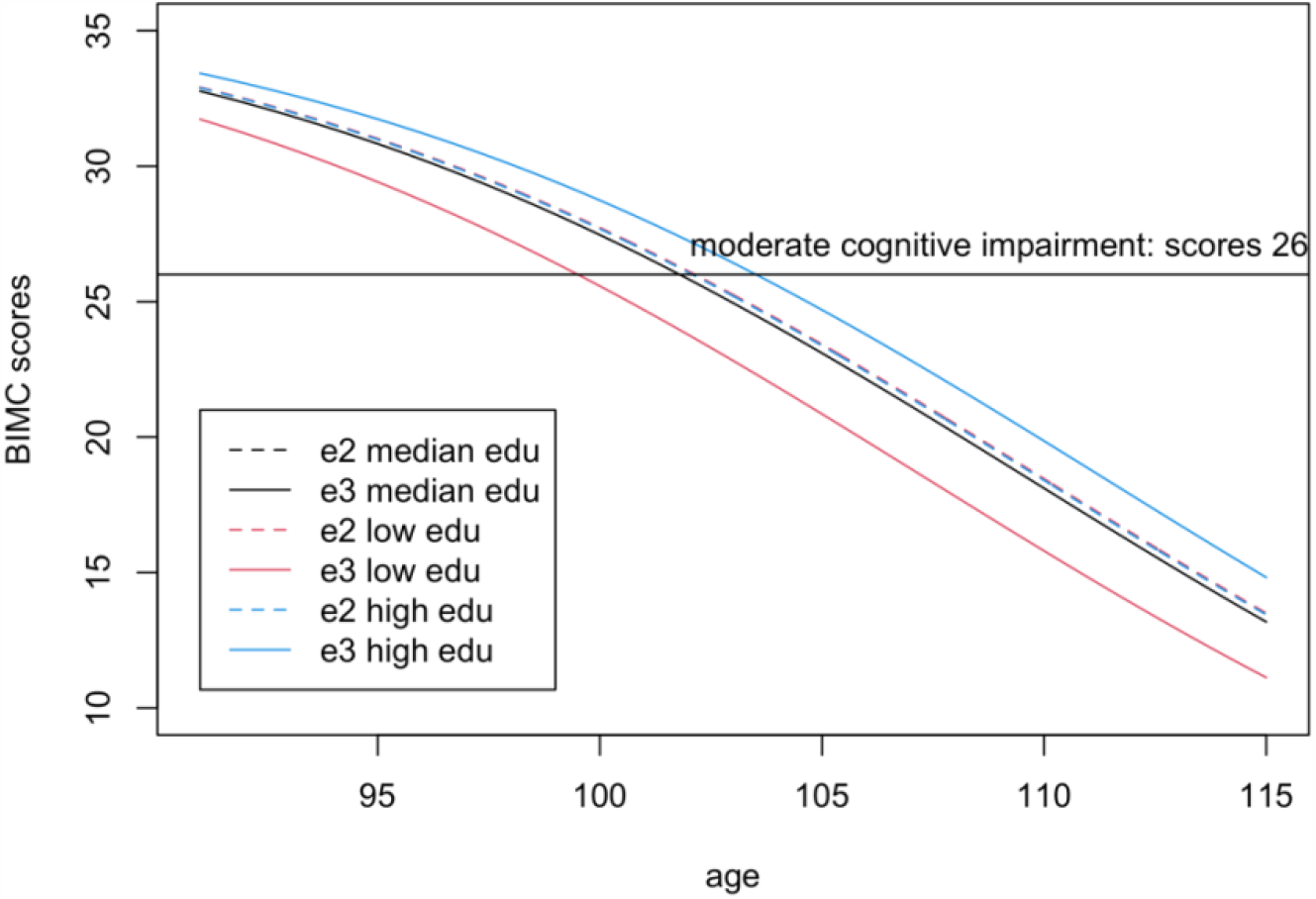
Predicted Blessed Information-Memory-Concentration scores in carriers of one or more e2 alleles (dashed lines) compared to homozygotes e3e3 (continuous lines) for low education (25% quantile, 8 years, red), median education level (12 years, black), and high education level (75% quantile, 15 years, blue). The scores were predicted using the estimates from Bayesian beta regression, assuming sex = female. Note visually there is only one dashed line, because the fitted lines of e2 carriers overlap in all three education groups.

Interestingly, only participants of the e3 group with low education had a worse BIMC score compared to carriers of one or more e2 alleles. The difference between the e2 and e3 groups became negligible in participants with median education level, and carriers of the e3e3 genotypes with a high level of education obtained a better score than carriers of the e2 alleles.

To better summarize the clinical implication of these results, we calculated the ages of onset of cognitive impairment predicted by the fitted model. From the fitted lines, we estimated that the ages of onset of moderate cognitive impairment were 99.5, 101.8 and 103.5 years in for e3 carriers with low, median and high education groups, respectively. The age of onset of moderate cognitive impairment (BIMC score = 26) was 102.1 years in e2 carriers, independent of education. Therefore, e2 carriers were estimated to delay the onset of moderate cognitive impairment by approximate 2 years compared to e3 carriers with low education, but this advantage essentially disappears with higher education.

In the analysis comparing e4 with e3, both the main effect term of e4 and the interaction with education were significantly negative, thus suggesting that higher education was not sufficient to remove the negative association of the e4 allele with BIMC scores. This is illustrated in Figure 3 that shows the predicted BIMC scores in the e3 and e4 groups stratified by education. Among individuals with the e3e3 genotype, more years of education was associated with higher BIMC scores, but the positive effect modification of higher education was reduced in carriers of e4 compared to e3.

**Figure 3.**
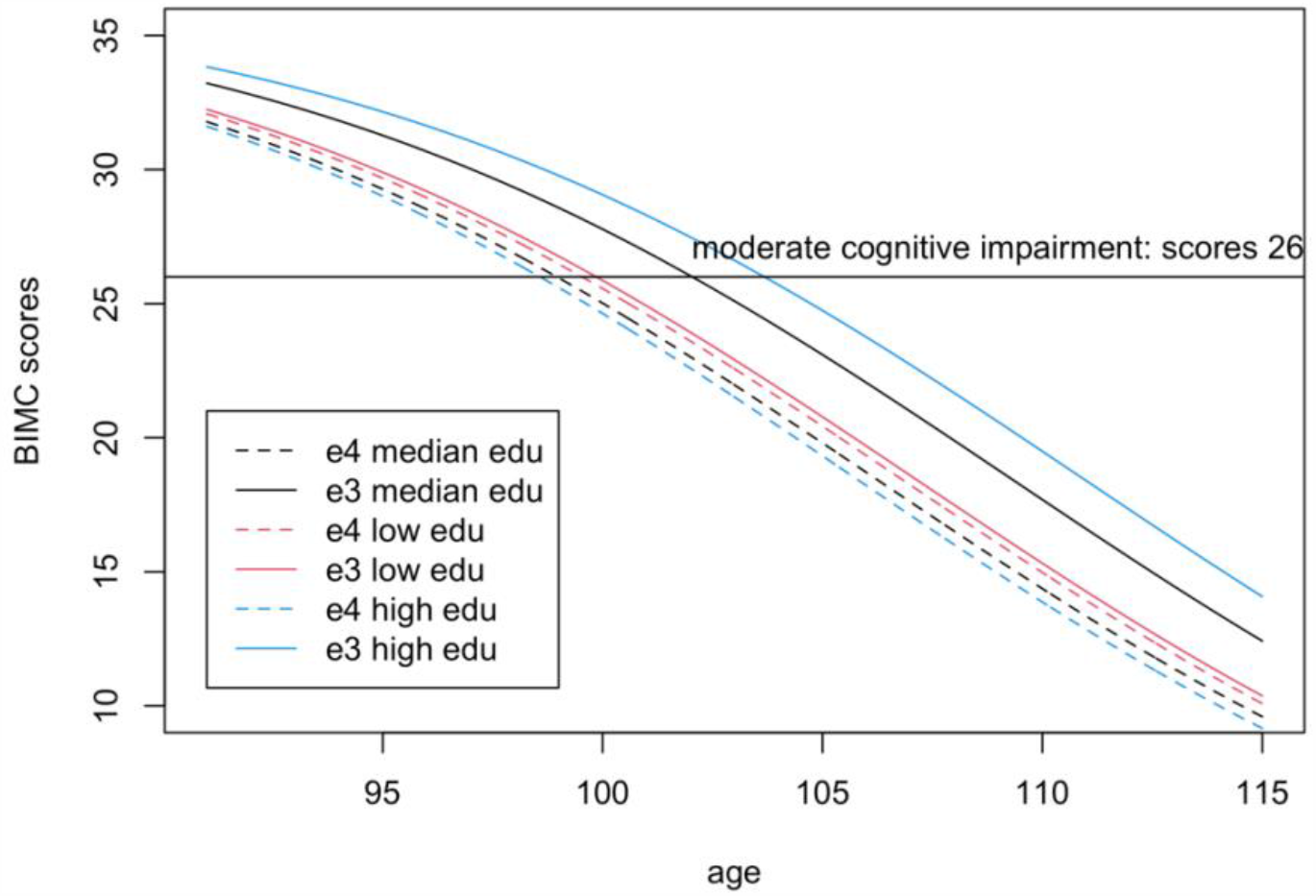
Predicted Blessed Information-Memory-Concentration test scores in carriers of one or more e4 alleles (dashed lines) compared to homozygotes e3e3 (continuous lines) for low education (25% quantile, 8 years, red), median education level (12 years, black), and high education level (75% quantile, 15 years, blue). The scores were predicted using the estimates from Bayesian beta regression, assuming sex = female.

From the fitted lines, the ages of onset of moderate cognitive impairment in e4 carriers were 99.5, 98.9 and 98.57 years in participants with low, median and high education respectively. Hence, in centenarians carrying the e4 allele, more years of education was not associated with a delay of cognitive decline.

## 4 Discussion

In this paper, we investigated the relationship between *APOE* alleles and change of cognitive function in a large cohort of centenarians enriched for carriers of the e2 allele. We used a Bayesian hierarchical beta regression model to characterize the association of *APOE* with cognitive function assessed through the Blessed Information-Memory-Concentration test. This method allowed us to fit the test scores in the range of admissible values, and to model non-linear relations of the score with age and education. Our analyses confirmed the decline of cognitive function and the positive association of education with preservation of cognitive function at extreme old age. The analysis also showed that the allele e4 has a negative association with cognitive function even at extreme old ages, while the allele e2 appears to be protective only in centenarians with low education. On the other hand, higher education appeared to increase the magnitude of the association between the e4 alleles and poorer cognitive function.

Our findings are consistent with previous studies that showed a decline of cognitive function with older age in multiple domains (Harada et al. 2013), and the importance of early education on better cognitive function at older age (Alley et al. 2007). Longitudinal studies of cognitive function in a large cohort of centenarians are limited. A study of Japanese centenarians detected an education by *APOE* e4 interaction on cognition that differed by sex (Ishioka et al. 2016). The study showed that highly educated centenarians who carried at least one e4 allele had worse performance in the Mini-Mental State Examination, compared to poorly educated centenarians. Our results are consistent with this counter-intuitive result but go one step further and disentangle the negative association of the e4 allele from the positive association of the e2 allele.

There is some evidence that the e2 allele confers protection from cognitive decline (Kim et al. 2017), in addition to increasing the chance for longevity (Sebastiani, Gurinovich, et al. 2019) and conferring protection from aging-related diseases (Wolters et al. 2019; Kuo et al. 2020). For example, we identified a cluster of participants in the Long Life Family study who have slower decline of processing speed and attention and are genetically enriched for the e2 allele (Sebastiani et al. 2020). These results suggest that targeting e2 gene products could lead to important therapeutics (Sebastiani, Monti, et al. 2019). The interaction between education and the e2 allele is also consistent with the fact that more years of education is associated with better cognitive function in later life (Alley et al., 2007; Wilson et al., 2009). Given the heterogeneity of the association between the e2 allele and longevity in populations with different genetic backgrounds (Gurinovich et al. 2019), it would be interesting to extend these analyses to also include the effect of nutrition on preservation of cognitive function as a possible effect modifier of *APOE* alleles.

The method we used in this analysis has several strengths. First, the cognitive test scores are bounded and skewed. Instead of applying an inadequate regression model assuming normal errors, we used Bayesian beta regression as a more appropriate modeling framework. Second, to increase statistical power, we included all participants with at least one cognitive function assessment in our model. A piecewise random intercept was used to include subjects with either repeated measurements or just one measurement. Finally, we carried out a model selection using the DIC criterion and then refined our model by credible intervals (CI). From the diagnostic plots of the MCMC, the final models converged well. Finally, in previous studies (Kulminski et al. 2015; Barral et al. 2017) *APOE* genotype was characterized as carriers of e4 (i.e. e2e4, e3e4, and e4e4), and non-carriers of e4 (i.e. e2e2, e2e3, and e3e3). This comparison could yield biased results since the non-carriers of e4 group might not represent the most prevalent population. To eliminate this potential bias, we divided the participants into subgroups, where we only compared the e2 carriers to e3 carriers, and the e4 carriers to e3 carriers.

Notable limitations of this study include loss to follow-up and mortality. Due to the extreme age of the study population, the mortality rate is considerably high: 49% of participants included in our analysis only completed the baseline evaluation. The absence of longitudinal data may induce bias, and thus we used a piecewise random intercept to compensate for those only with one record.

In conclusion, we examined the association between *APOE* alleles and cognitive decline in a cohort of centenarians. We confirmed the negative correlation between the e4 allele and cognition even at the extreme of human lifespan, and newly found the carrying the e2 alleles appears to be beneficial only in centenarians with poor education. The interaction between *APOE* e4 and education produced a counter-intuitive result that is however consistent with other results in centenarians. The antagonistic relation between higher education and carrying the e4 allele may be confounded by other risk factor and warrants more in depth studies.

## Data Availability

The data is restricted and available upon request.

## 6 Author Contributions

Author Q. X. proposed the method used in the paper and conduct the statistical analysis and drafted the manuscript.

Author S. A. and T. P. designed the study and collect data from study cohort and helped editing the manuscript.

Author P. S. designed the analytic strategy, supervised the work and critically revised the manuscript.

